# Mortality of adults with chronic noncancer pain: a systematic review and meta-analysis

**DOI:** 10.1101/2024.03.22.24304748

**Authors:** Stephanie Webb, Antonia-Olivia Roberts, Lauren Scullion, Georgia C. Richards

## Abstract

It is recognised that chronic pain is one of the leading causes of disabilities worldwide. However, statistics on mortality and causes of death in people with chronic noncancer pain (CNCP) have been difficult to determine. This systematic review aimed to determine the mortality rate in people with all types of CNCP and the associated causes and risk factors of death. MEDLINE (Ovid) and EMBASE (Ovid) were searched on 23 March 2023 to identify epidemiological studies reporting mortality in people with CNCP. Nineteen observational studies were included. There were 28,740 deaths (7%) reported in a population of 438,593 people with CNCP (n=16 studies), giving a mortality rate of 6,553 deaths per 100,000 people. An exploratory meta-analysis found that the relationship between mortality and CNCP was statistically significant (mortality risk ratio: 1.47; 95% CI: 1.22-1.77; n=11 studies) when comparing people with CNCP to those without pain. People with CNCP were more likely to die from cardiovascular disease whereas those without pain were more likely to die from malignancy, respiratory and gastrointestinal diseases. Smoking, lower physical activity levels, and opioid use were risk factors for death in people with CNCP. This systematic review found that people with CNCP have a higher risk of mortality than people without chronic pain. To reduce mortality rates in people with CNCP, cardiovascular diseases and risk factors for death should be considered when managing people with CNCP.

## 1 Introduction

Chronic pain and pain-related diseases are the main cause of years lived with disability (YLD) and disease burden globally [15,16]. In the United States (US), the Centre for Disease Control (CDC) estimated that one in five adults live with chronic pain [8]. In the United Kingdom (UK), a systematic review of observational studies estimated that one in two to one in three adults live with chronic pain [14]. After onset, chronic pain persists [27], which negatively impacts the person’s quality of life [10] and the healthcare systems in which they live [26]. Despite the widespread impact of chronic pain, data on the mortality of these individuals is often poorly reported and unexplored.

The World Health Organisation (WHO) updated the International Classification of Diseases (ICD) in 2019 (i.e. ICD-11) to capture chronic pain subtypes [47] including widespread, localised, primary, secondary, and cancer pain [47]. Despite the new ICD codes, public health authorities are yet to routinely capture and monitor mortality statistics in people with chronic pain. For example, the National Health Service (NHS) in England will not mandate the use of ICD-11 codes until April 2026 [28]. While these statistics are difficult to obtain, the number of deaths in people with chronic pain have increased due to the ubiquitous use of opioids [42]. In contrast, statistics on people with cancer are well documented. In 2018, 149.2 people in every 100,000 died of cancer in the US, [5] while in England and Wales, 266 people died per 100,000 [33] In people with cancer, pain is reported in 38% to 56% of cases [3,4].

A systematic review of ten observational studies of people with all types of pain was published in 2014 [38]. This review found a modest but non-significant increase in mortality in people with chronic pain (mortality rate ratio: 1.14; 95% CI 0.95–1.37) [38]. However, this review included people with cancer pain and did not explicitly focus on chronic pain. In 2017, a systematic review of all-cause mortality in people with chronic widespread pain was published which included cancer-related mortalities [21]. This review found a pooled estimate of all-cause mortality in people with chronic widespread pain to be 57% (1.57) but had significant heterogeneity. There were two registered systematic reviews investigating mortality in people with spinal pain [32] and back pain [35]. However, no review has focused on mortality in people with chronic non-cancer pain (CNCP) and the associated risk factors. Therefore, this systematic review aimed to determine the mortality rate and associated risk factors in people with all types of CNCP excluding cancer-related pain.

## 2 Methods

The protocol for this systematic review was preregistered on an open repository (https://doi.org/10.17605/OSF.IO/7E48V) [45].

### 2.1 Searches

MEDLINE (Ovid) and EMBASE (Ovid) were searched on 23 March 2023 using keywords relating to “chronic pain”, “chronic noncancer pain”, “death”, and “mortality”. The search strategy is in Supplement Table S1 and S2.

### 2.2 Screening

Two researchers (SW (all studies) and AR or LS as second reviewers) independently screened articles for their eligibility, blinded to each other’s decisions. A third reviewer (GCR) was consulted if there were discrepancies in the two screening stages: title and abstracts and full-text screening. Rayyan software was used to complete the screening.

Studies were included if they involved (1) all humans of all ages who were clinically diagnosed with CNCP, which was defined as pain for 12 or more weeks (i.e., three or more months) or coded using ICD-11 [42]; (2) data on death; (3) were observational studies, including cohort, case-control, and cross-sectional studies; (4) cancer as a cause of mortality associated with pre-existing CNCP; and (5) were published in the English language.

Studies were excluded if they involved animals or non-human participants; involved people with acute pain (i.e., pain for less than 12 weeks) or cancer pain; and were clinical trials, case reports, test-tube experiments, commentaries, editorials, letters, or systematic reviews.

### 2.3 Data Extraction

Data were extracted into a pre-defined data extraction template using Google Sheets by one researcher (SW) and cross-checked by a second reviewer (GCR). The following variables were extracted: Study ID (Surname of first author, year of publication), title, journal, aims, population characteristics (including the number of participants, age, gender or sex, location, ethnicity, and pain phenotype), methods (including when the study was conducted, study design and statistical analyses used, follow up time), results (including the number of deaths, all-cause mortality, cause-specific mortality, causes of death and risk factors associated with deaths if reported).

### 2.4 Quality Assessment

The quality of studies was critically appraised by one researcher (SW) using the National Institute of Health (NIH), National Heart, Lung, and Blood Institute (NHLBI) Quality Assessment Tool for Observational Cohort and Cross-Sectional Studies [29] This tool was selected as it accounted for both cohort and cross-sectional studies simultaneously. There were 14 domains evaluated by this tool to assess the quality of the research question: the reporting of the study population, participation rate, selection of participants, sample size, appropriateness of statistical analyses, timeframe for associations, levels of exposures, ascertainment of the exposure, appropriateness of outcome measures, outcome blinding of assessors, loss to follow-up, and adjustment for confounding. For the 14 criteria assessed, a score of “fair” was assigned if the authors did not adequately justify their sample sizes or did not report whether participants were lost to follow-up and did not control for confounding variables, studies that adequately met these criteria were rated as “Good”.

### 2.5 Data analysis

One researcher (SW) descriptively analysed the extracted data using percentages for categorical variables and median and interquartile ranges (IQR) for continuous variables. The risk ratios for mortality reported in studies were given as ranges for each type of ratio. The total number of participants with chronic pain was summed across all studies, as well as the number of deaths reported to calculate the mortality rate. The total number of deaths by specific cause were summed across studies, as were those that included a comparator no pain group. For the deaths by cause, the percentage of deaths by cause was calculated by dividing by the total deaths and presented graphically.

Studies that had a chronic pain and a no pain group (or control group) were included in the exploratory meta-analysis using the Mantel-Haenszel method with a random effects model to determine the overall risk ratio for mortality using RevMan [41]. Sensitivity analyses were conducted to assess heterogeneity by grouping studies by sample size, data source, pain definition, country of origin, decade of publication and decade of study commencement. Factors associated with mortality and their method of determination were narratively combined and reported as frequencies.

### 2.6 Software and data sharing

Several different software packages were used to conduct this review, including Mendeley to remove duplicates, Rayyan to screen studies for eligibility, Google Sheets to extract data and for descriptive analyses, and RevMan [41] to conduct the exploratory meta-analysis. The study protocol, data and study materials are all available on an open repository [45] (https://osf.io/cp3fx/?view_only=09efe9b9b60e4d81950f248cb08f6867).

## 3 Results

Nineteen studies [1,2,7,9,11,12,18,19,21–25,30,36,37,43,44,46] met the eligibility criteria and were included in this systematic review (Figure 1). During full-text screening, 73 studies were excluded as they: did not meet the definition for chronic pain (n=39), were the wrong publication type (n=12), had the wrong outcome (n=10), included data on malignant pain (n=5), were the wrong study design (n=8), and one study was published in Spanish.

**Figure 1:**
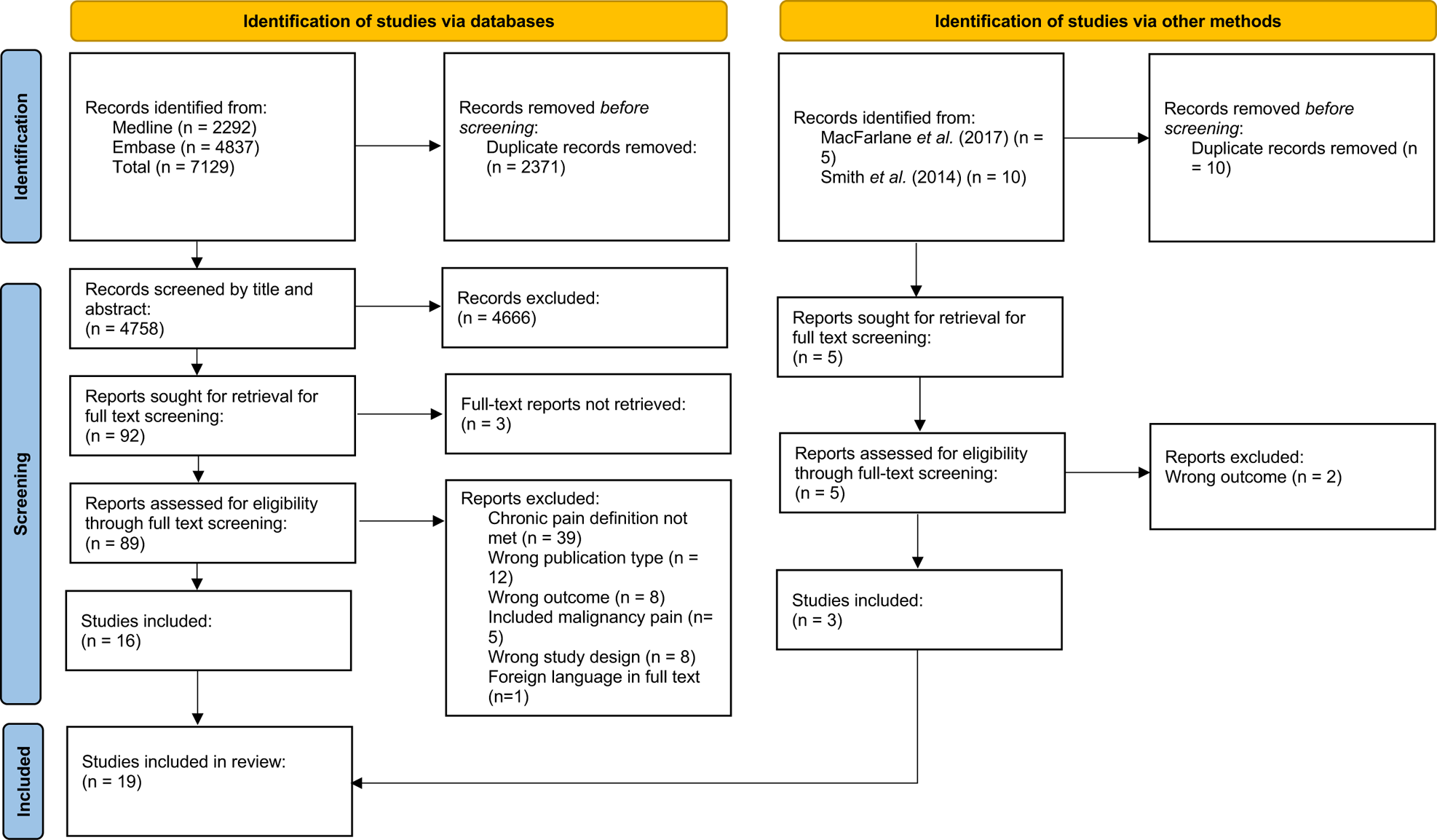
PRISMA flow diagram of the screening of studies for eligibility in this review.

The 19 included studies were all prospective cohort studies and enrolled a combined total of 443,239 participants with CNCP. These participants were not evenly distributed across the 19 studies, with a median of 2,168 participants per study (range: 249-187,269) (Table 1). The majority of participants were recruited from the UK Biobank [7,21,22], followed by surveys [1,2,11,12,18,19,23,25,30,36,37,44], in-patient clinics [9,24,43], and other databanks [46]. Most studies were conducted in the United Kingdom (37%; n=7), followed by Denmark (21%; n=4) and the United States (US; 21%; n=4), Norway (11%; n=2) and Sweden (11%; n=2). Most participants were female (mean 64%; 95% CI: 57-71%).

**Table 1:**
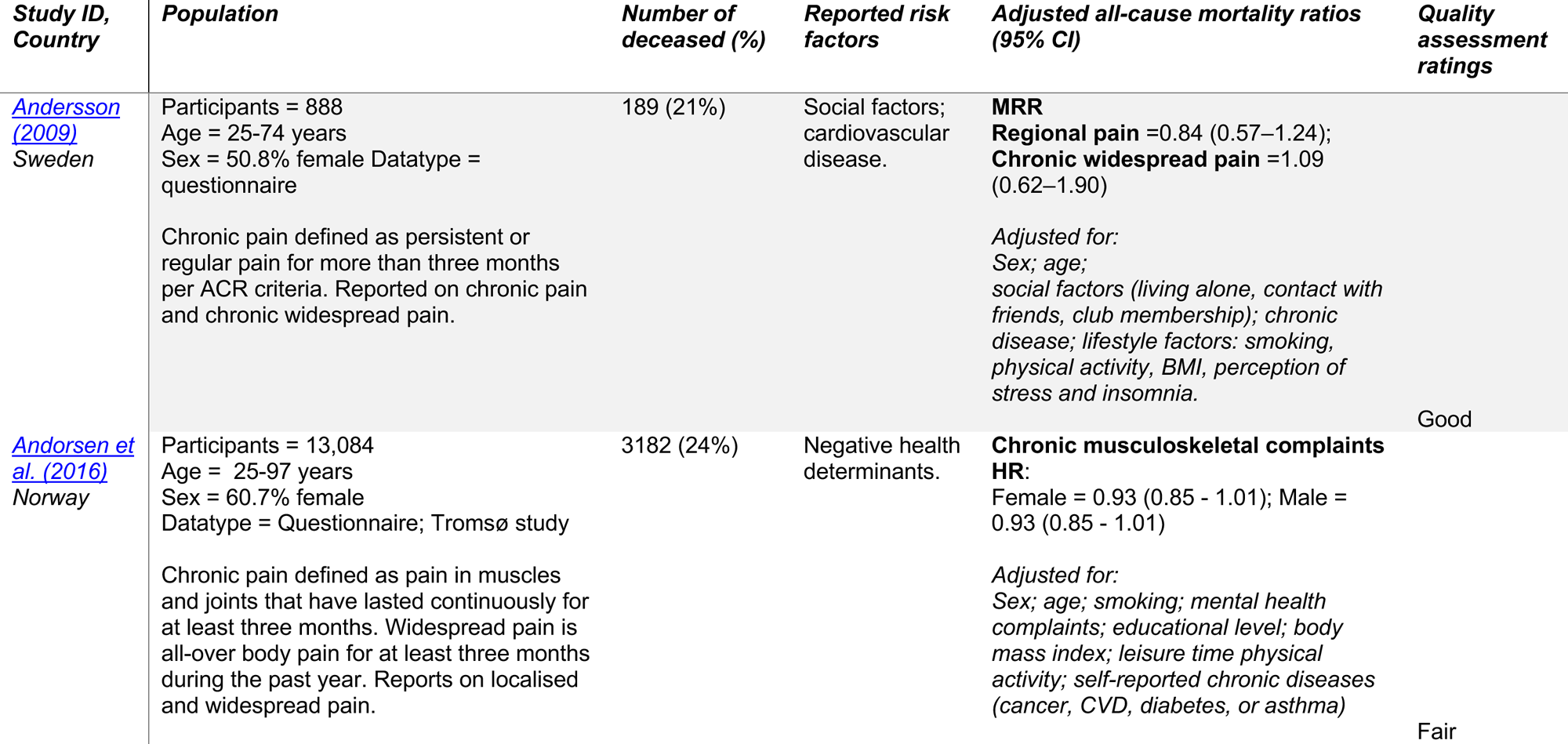
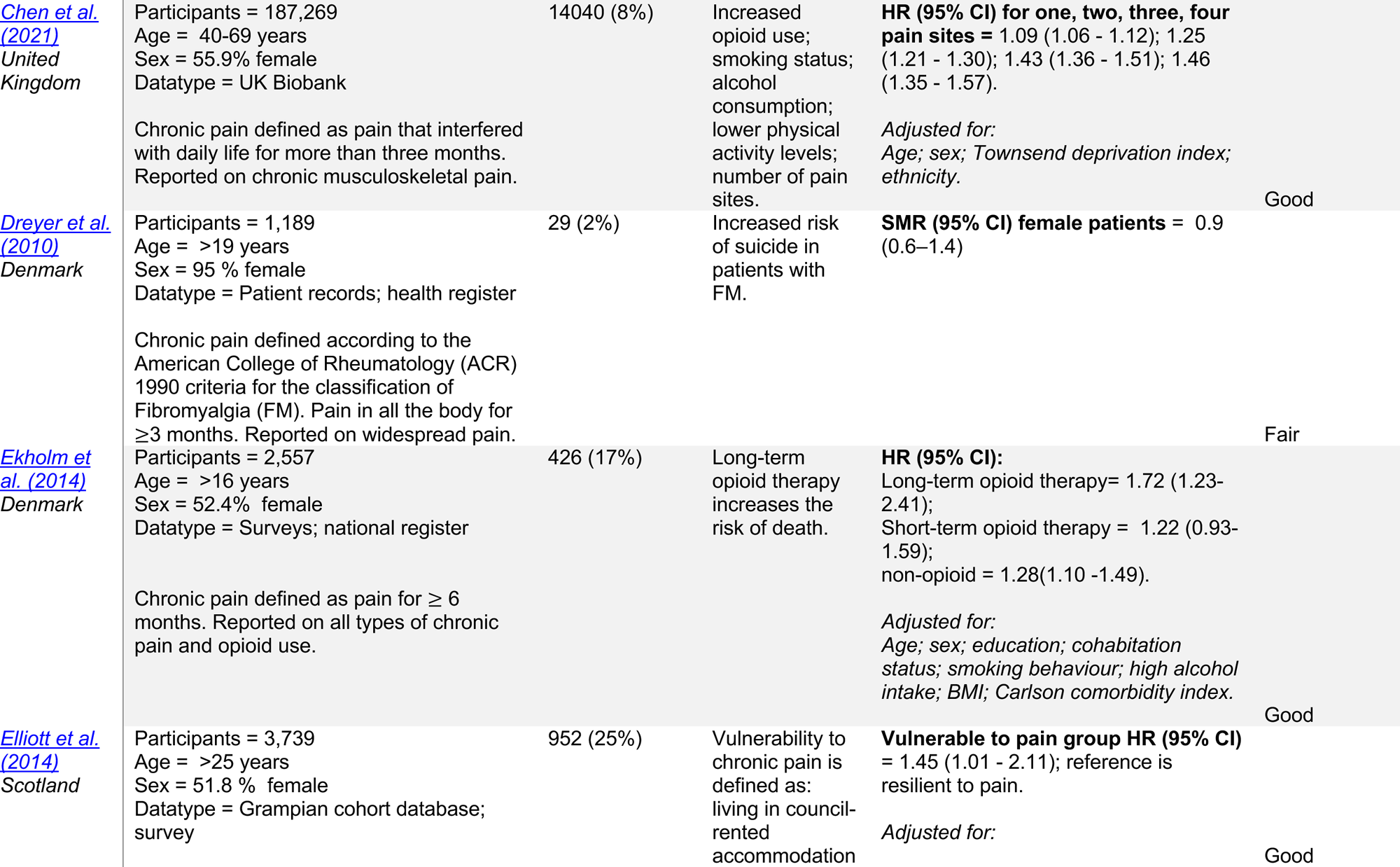
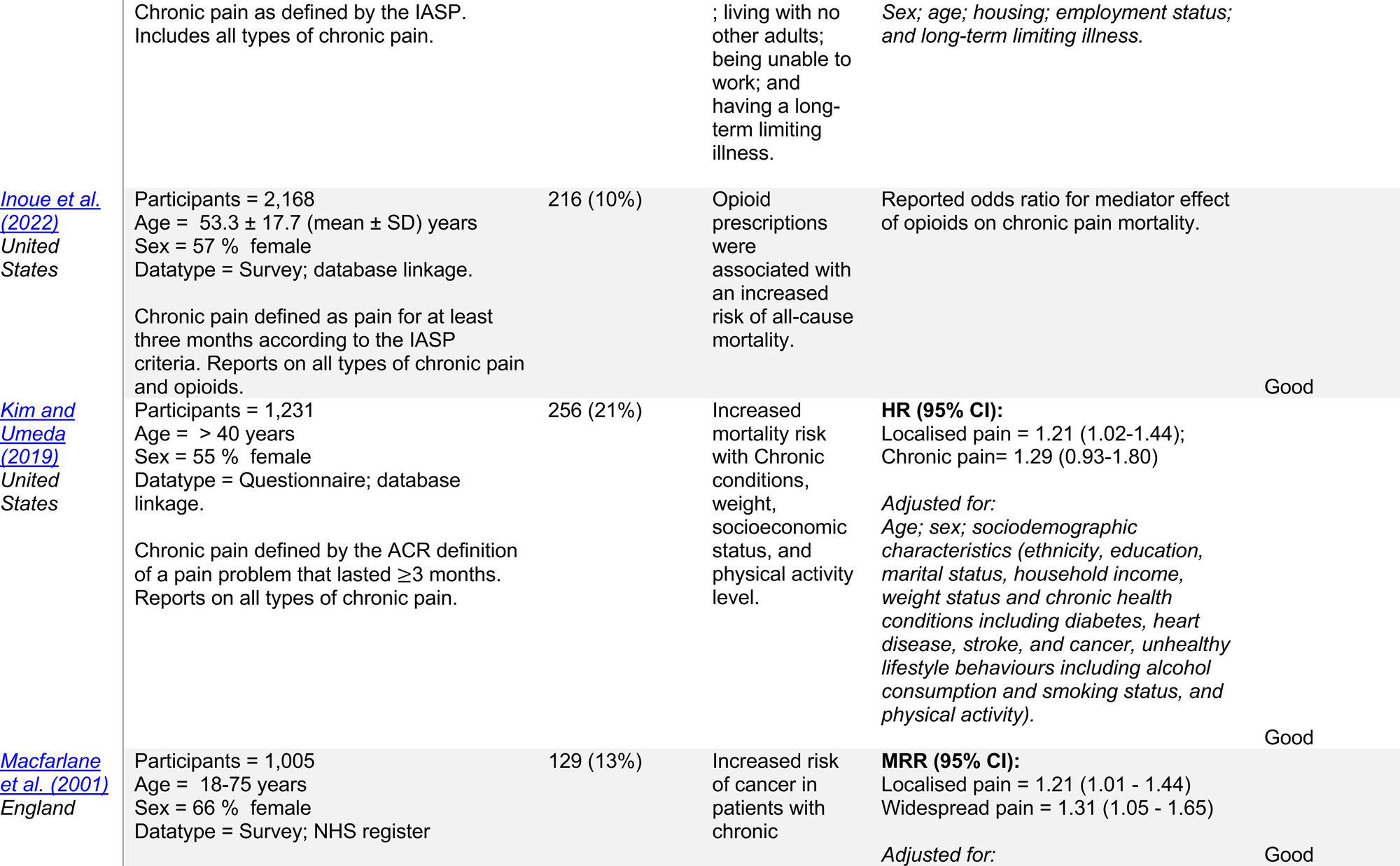
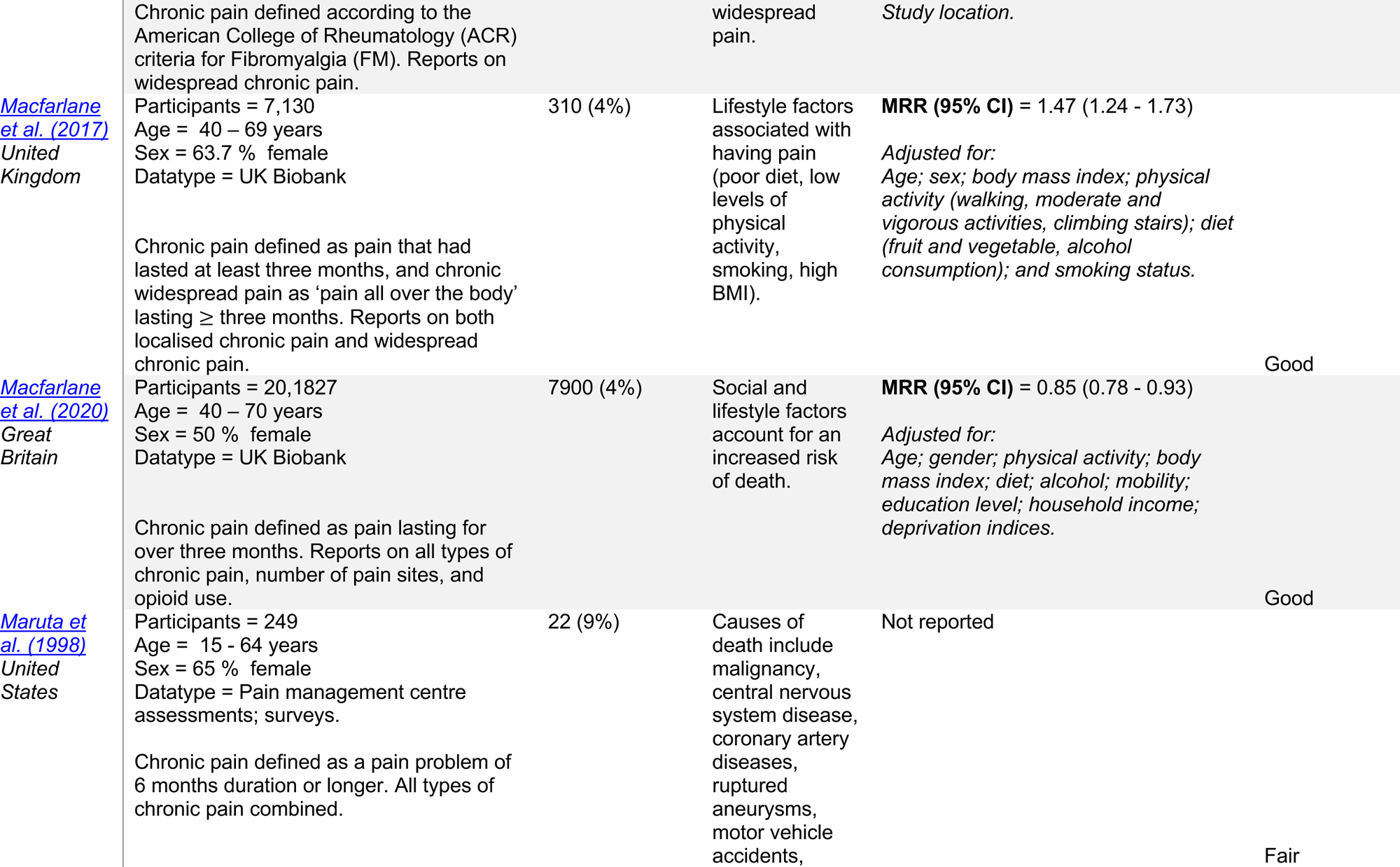
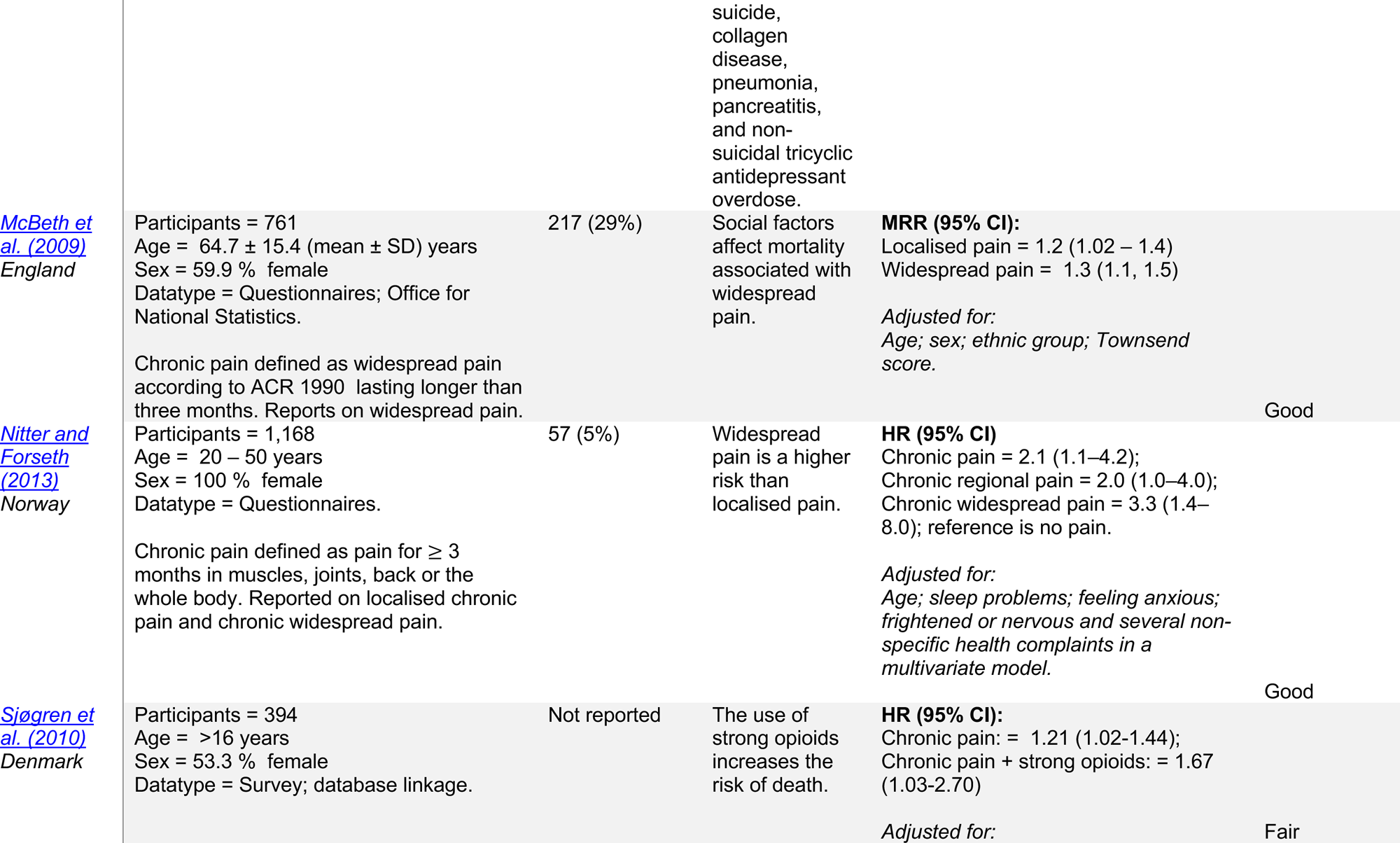
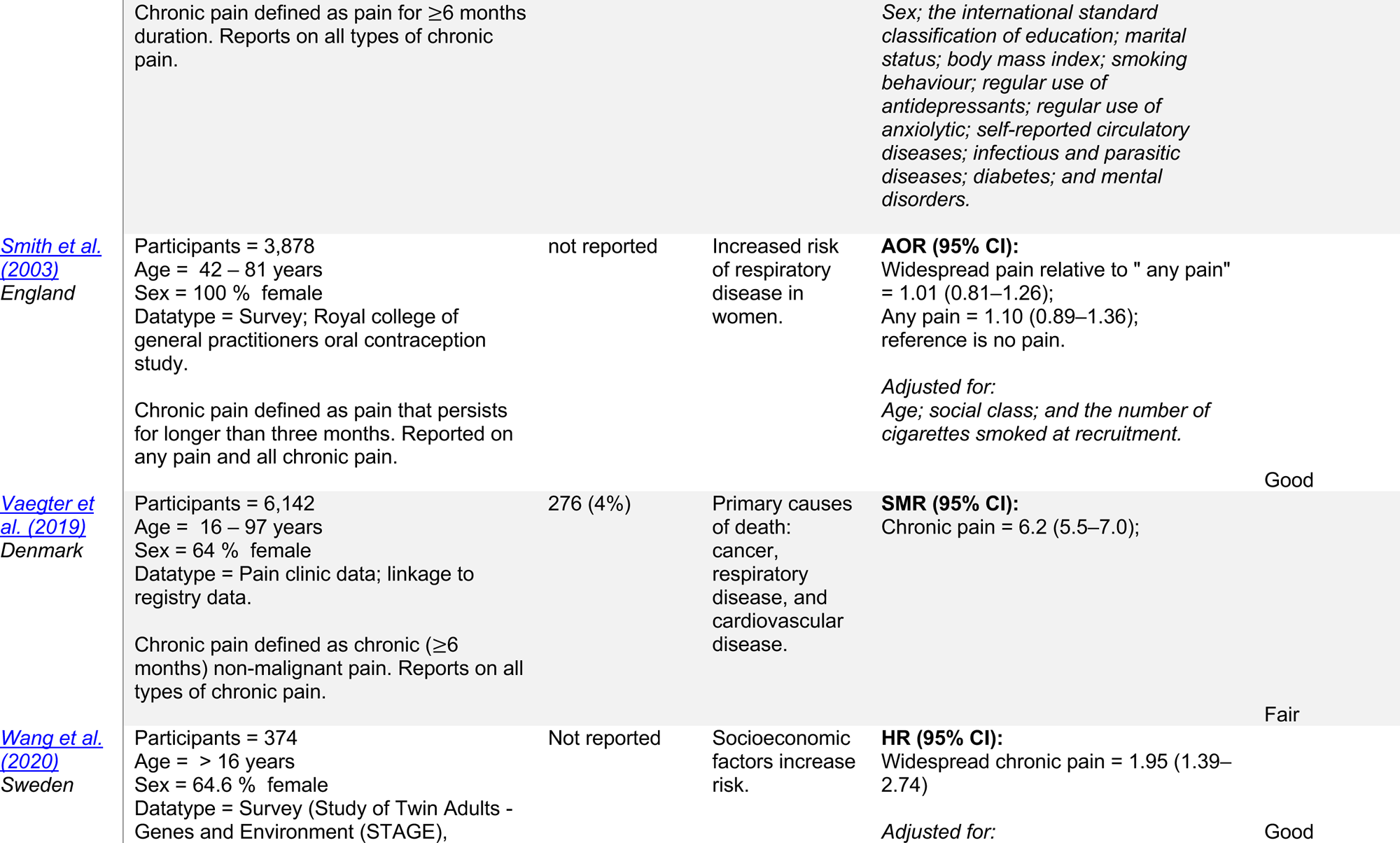
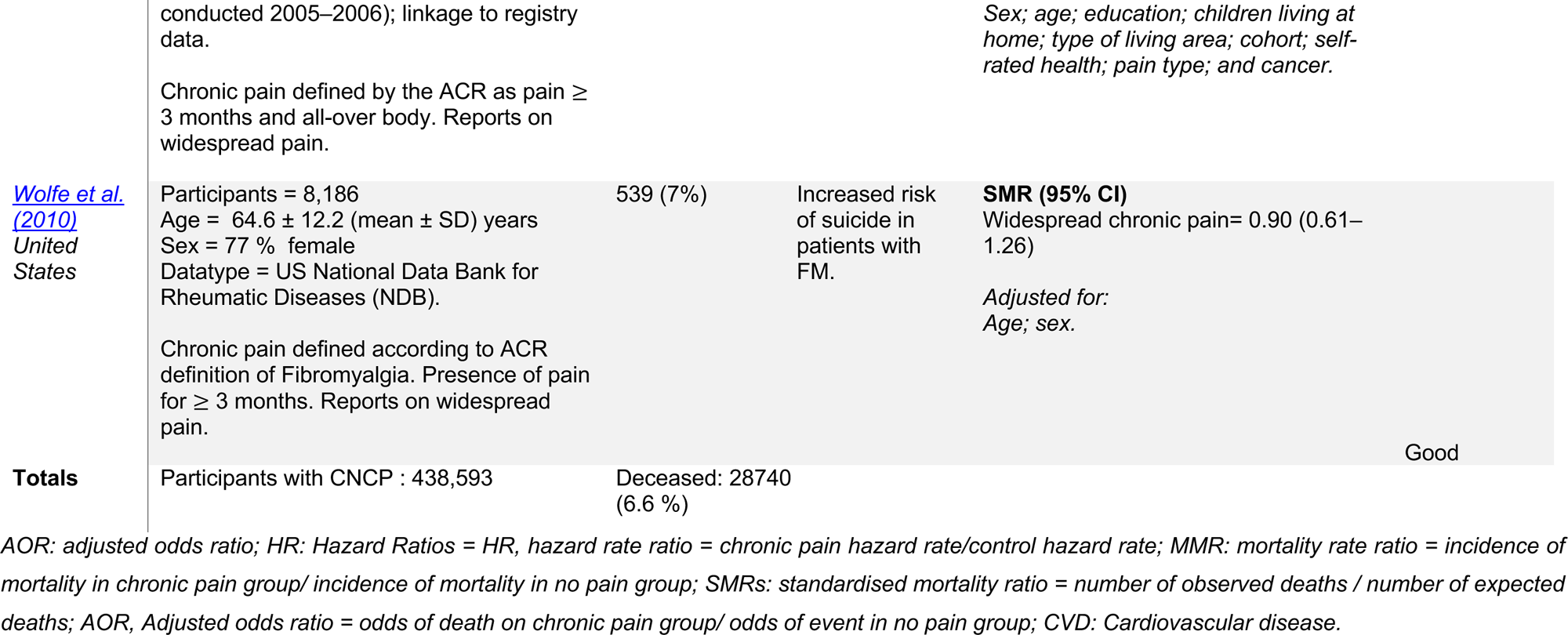
Summary of study characteristics from the 19 included studies on mortality in participants with chronic non-cancer pain, including reported risk mortality ratios and the quality assessments of the studies using the National Institute of Health (NIH), National Heart, Lung, and Blood Institute (NHLBI) Quality Assessment Tool for Observational Cohort and Cross-Sectional Studies.

To define chronic pain, six studies stated that chronic pain was pain ≥ 3 months duration [2,7,21,22,30,37], seven studies used the American College of Rheumatology (ACR) criteria [1,9,19,23,25,44,46], four defined chronic pain as pain that persisted for ≥ 6 months [11,24,36,43], and two referred to the International study for the Association of Pain (IASP) definition [12,18]. The types of CNCP also varied, with five studies reporting both localised and widespread pain [1,2,21,22,30], five studies reported widespread pain [9,23,25,44,46] three specifically referred to fibromyalgia [9,23,46] one study focussed on chronic musculoskeletal pain [7], and nine studies did not specify CNCP subgroups [11,12,18,19,22,24,36,37,43].

### 3.1 Mortality in people with chronic pain

Of the 19 included studies, 16 (84%) reported raw data on the number of deaths, where 438,593 participants had CNCP of which 28,740 deaths were reported, equating to a mortality rate of 6.6 deaths per 100 people with CNCP (Table 1).

Seventeen (90%) studies reported various ratios, with nine (53%) studies reporting hazard ratios (HR) [2,7,11,12,19,30,36,44], five (29%) studies reporting mortality risk ratios (MRRs) [1,21–23,25] three (18%) studies reported standardised mortality ratios (SMR) [9,43,46], and one study reported the adjusted odds ratio (AOR) [37]. These ratios were adjusted for various population characteristics (Table 1). For any type of CNCP, the ranges of the reported ratios were HR: 0.93–2.1; MRR: 0.84–1.47; and SMR: 0.9–6.2. Two studies did not report any ratios for mortality [18,24] Inoue *et al.* reported the effect of opioids on mortality in people with chronic pain using odds ratios (OR) [18], and Maruta *et al*. reported on causes of death [24].

Eleven studies [1,2,7,11,18,19,21–23,25,30] included a reference or control group that reported no pain (Table S3 in supplement). In this cohort of 11 studies, there were 419,088 participants with CNCP of which 26,931 deaths were reported, equating to a mortality rate of 6.4 deaths per 100 people. There were 786,626 participants without CNCP of which 35,033 deaths were reported, totalling 4.5 deaths per 100 people. Those with CNCP had a 30% greater mortality rate than those with no CNCP.

### 3.2 Exploratory meta-analysis

To further explore the differences in mortality between participants with and without CNCP, a meta-analysis was conducted using data from the eleven studies with comparison groups. This meta-analysis found a statistically significant risk ratio of 1.47 (95% CI: 1.22 - 1.77) (Figure 2). However, there was a high degree of heterogeneity (I^2^ = 99%). To examine the drivers of the heterogeneity several sensitivity analyses were conducted, including grouping studies by sample size, the source of data, study start date, study publication date, definition of pain and country of origin. However, the heterogeneity persisted (see Supplement Figures S6-S11).

**Figure 2:**
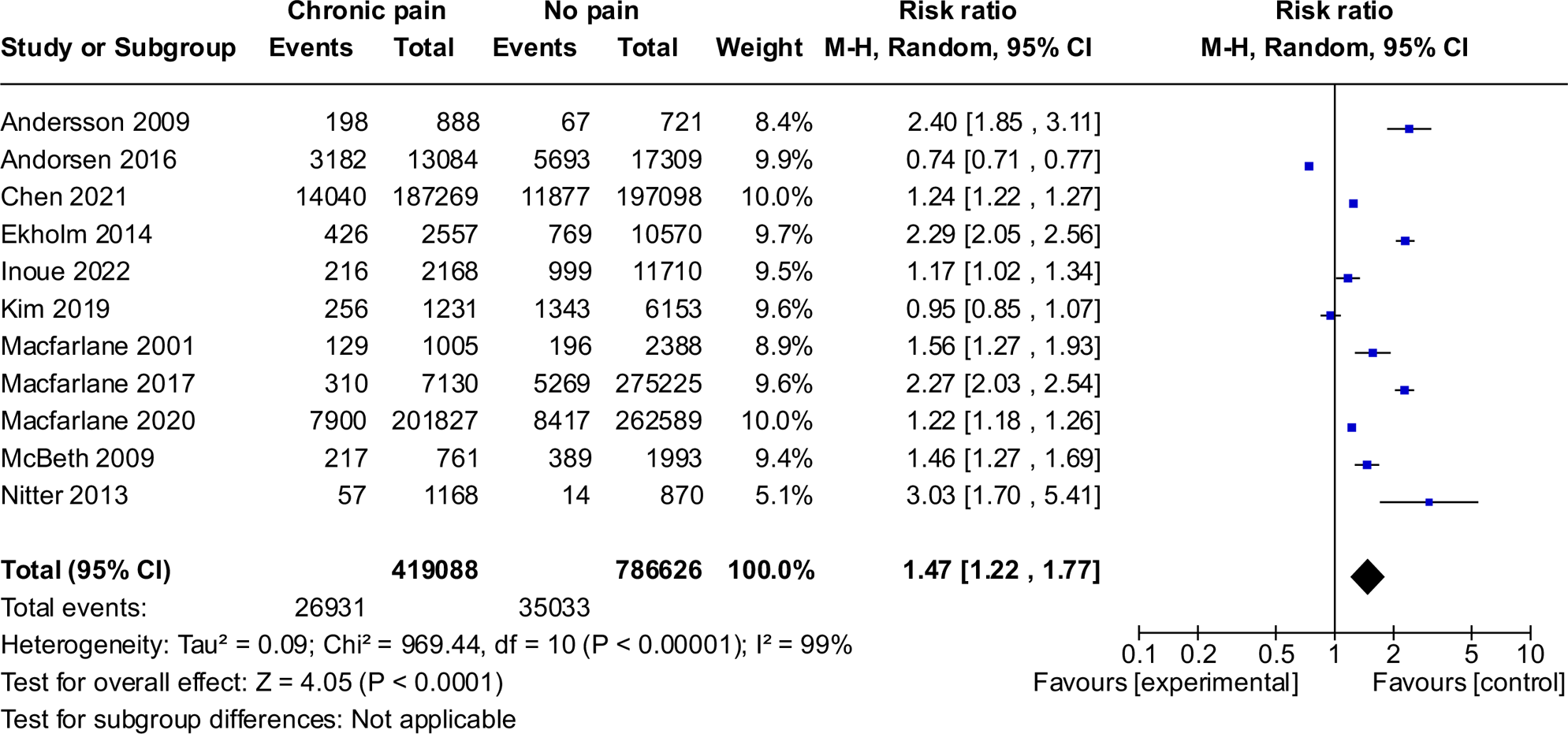
Exploratory meta-analysis of the 11 studies that included deaths for those with and without chronic noncancer pain. Events; number of deaths reported in study. Total; number of participants included in studies.

### 3.3 Causes of death and risk factors

Of the 19 included studies, 10 studies (53%) [1,2,7,9,12,23–25,30,43,46] reported the number of deaths by specific cause in the CNCP population, totalling 15,108 deaths. Of these causes of death, people with CNCP died most often from malignancy (n=8511); cardiovascular diseases (CVD) (n= 4679); nervous system diseases (n= 764); and mental health disorders (n= 317) (Table S4, Supplement).

Of the 10 studies that reported specific causes of death, three (16%) reported [1,7,30] causes of death in a non-pain comparator group, totalling 11,004 deaths. The main causes of death for the non-pain population were malignancy (n= 6,533), CVD (n= 2,274), respiratory disease (n = 720) and nervous system disease (n = 649). Comparing the causes of death in the CNCP and non-pain groups (Figure 3), death from malignancy was similar across these populations. Deaths from CVD and suicides were higher in the CNCP population, whereas deaths from respiratory and gastrointestinal diseases were relatively more common in the non-pain group.

**Figure 3:**
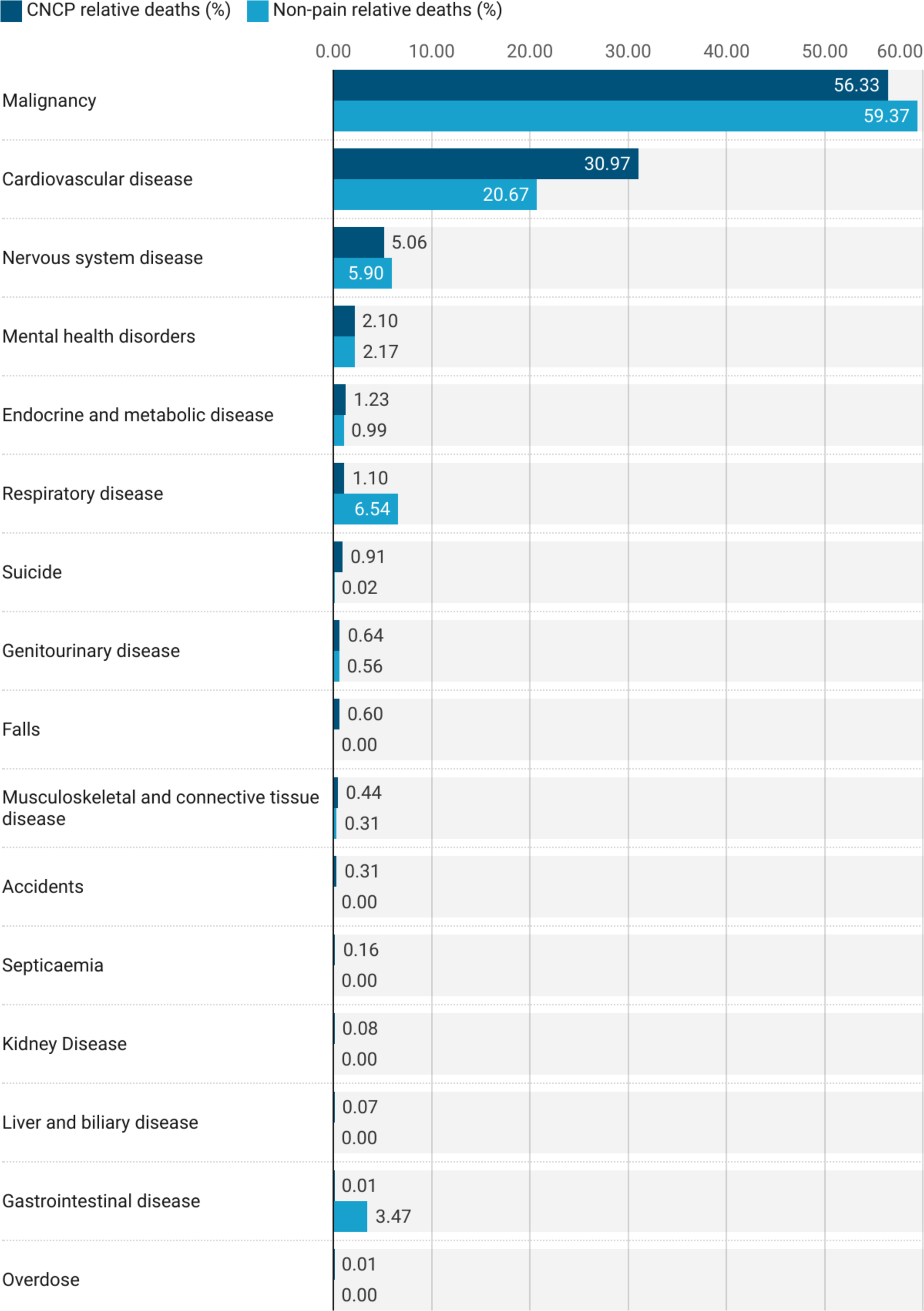
Frequency of deaths by cause relative to total number of deaths reported for the chronic noncancer pain (CNCP) and non-pain groups. For CNCP there were 15,108 deaths reported by cause and for non-pain there were 11,004 deaths reported by cause. The deaths are described as a percentage of these reported deaths.

In total, 12 studies (63%) [1,7,11,12,18,19,21,22,30,36,44,46] reported risk factors for mortality in people with CNCP. Of these studies, six used covariate analysis (31%) to assess the effect on risk ratios [1,12,21,22,44,46], finding a decrease in risk ratios when controlling for these variables. Five studies (26%) assessed risk factors directly by collecting data on these variables [11,19,22,30,36], and two studies (10%) assessed risk factors through mediator analysis [7,18].

The risk factors for mortality in those with CNCP were reported in studies as follows: smoking (n = 5; 26%)[1,7,19,21,22]; lower physical activity levels (n = 4; 21%) [1,19,21,22]; opioid use (n = 4; 21%) [7,11,18,22]; high body mass index and/or obesity (n = 3; 16%) [1,19,22]; alcohol consumption (n = 3; 16%) [7,19,22]; a higher number of pain sites or pain intensity (n = 3; 16%) [7,22,30]; having a long-term illness (n = 2; 10%) [12,19]; low income/higher deprivation (n = 2; 10%) [19,22]; socioeconomic factor (n = 2; 10%) [44,46]; housing status (n = 1; 5%) [12]; unemployment (n = 1; 5%) [12]; disability (n = 1; 5%) [12]; anxiety (n = 1; 5%) [30]; and educational attainment level (n = 1; 5%) [44].

### 3.4 Quality Assessment

Fourteen (74%) studies were rated “good” [1,7,11,12,18,19,21–23,25,30,37,44,46] and five [2,9,24,36,43] (26%) were rated “fair” (Table 1; Supplement Table S5). Of the studies rated fair one study did not provide a sample size justification or an effect estimate [24], two studies had a loss to follow-up of greater than 20% [2,36], and three studies did not control for confounding variables [9,24,43]. Two studies anonymised the data before analysis [9,43]. None of the included studies assessed the sample population’s chronic pain status more than once. Seven of the included studies assessed chronic pain using scales [1,2,7,19,22,23,30]. Nine studies reported loss to follow-up [1,9,12,18,24,25,30,43,46].

## 4 Discussion

People with CNCP had a 30% greater mortality rate than those without pain. For every 100 people with CNCP, between six and seven died in the included studies, while between four and five people died without chronic pain. Those with CNCP were more likely to die from cardiovascular disease (CVD) whereas those without pain were more likely to die from malignancy, respiratory, and gastrointestinal diseases. Smoking, lower physical activity levels, and opioid use were risk factors for death in people with CNCP.

### 4.1 Comparison to previous literature

In contrast to other systematic reviews that focused on chronic pain [38] or specifically widespread chronic pain [21], this review identified 19 studies and is generalisable to all types of CNCP, excluding pain related to malignancy.

Smith *et al.* [38] found a modest but non-significant increase in the risk of mortality in people with chronic pain from ten included studies. They conducted meta-analyses of seven included studies that resulted in a non-significant MRR of 1.14 (95% CI 0.95– 1.37; I^2^ = 79%). Macfarlane *et al.* [21] found a significant excess risk of mortality in people with chronic widespread pain with an overall MRR of 1.57 (95% CI 1.06-2.33; I^2^=98%; n=6 studies). We similarly found a significant risk ratio of 1.47 (95% CI 1.22-1.77; I^2^=99%; n=11 studies) in our exploratory meta-analysis of mortality in people with CNCP.

Macfarlane *et al.* [21] conducted a meta-analysis by cause of death and found that three out of five included studies showed excess mortality for CVD in people with widespread chronic pain in comparison to people with no pain. This result is concordant with our finding that CVD is a more common cause of death in those with CNCP. They also reported increased mortality rates for malignancy and respiratory disease death in people with chronic widespread pain. This result contrasts our finding that malignancy death rates are comparable between groups and respiratory disease deaths are higher in those with no pain. Meanwhile, Smith *et al.* [38] reported modest increases in the risk of mortality from CVD, respiratory disease and malignancy for people with chronic pain, but these values were non-significant. Although these meta-analyses report by cause of death they do not give the number of raw deaths, and report only studies that used MRR as an outcome measure.

There is evidence to support the link between CNCP and CVD [13,25,31,34,40]. A systematic review and meta-analysis found that people with chronic pain had an increased risk of CVD-related mortality, which found a link between pain intensity and CVD risk [13]. A second meta-analysis confirmed a similar relationship between musculoskeletal pain and CVD [31]. A cohort study using the UK Biobank found a statistically significant increased risk of CVD in patients with chronic pain when controlling for lifestyle factors [34]. Moreover, a cohort study found that those living with fibromyalgia have an increased risk of coronary heart disease [40].

The relationship between CNCP and CVD may be explained by the social and lifestyle factors (e.g. smoking and physical inactivity) that this review found were commonly reported as risk factors for increased mortality in people with CNCP. These risk factors have similarly been reported in a narrative review of lifestyle factors in people with chronic pain [26]. However, whether these lifestyle factors precede the chronic pain diagnoses or are a consequence of chronic pain is unknown. Therefore, prospective cohort studies should assess chronic pain status, lifestyle factors, and health behaviours at multiple time points alongside mortality and specific causes of death to evaluate this association.

The finding that respiratory disease is a more common cause of death for those without pain and that malignancy is similar between the CNCP and non-pain groups, is noteworthy, as diseases such as COPD are commonly comorbid with CVD [6] and malignancies [20]. Determining why these differences exist is important to explore for developing management regimens for patients living with CNCP.

This review found that the use of opioids in people with CNCP was a risk factor for mortality. The association between opioids and mortality in comparison to other pain management strategies in chronic pain patients has been assessed in other systematic reviews [39,42]. These reviews found that opioid substitution or alternative treatments reduced mortality in chronic pain populations. Further research into this subpopulation of chronic pain patients prescribed long-term and potentially high-dose opioids is required to understand the harms over time.

### 4.2 Strengths and limitations

This systematic review is the first to synthesise studies that exclusively focus on all types of CNCP, so the findings are generalisable to people who experience various chronic pain conditions, excluding malignant pain. However, the findings may not be generalisable to people with CNCP outside of Europe and North America as there were no studies published in these regions. Due to the growing global burden of chronic pain, future research should assess the mortality rates of those with CNCP in South America, Africa, and Asia to determine the global mortality rate. Although the included studies were observational, none were rated as ‘poor’ in the quality assessment, and all had a prospective design.

Across the studies, there were variations in the acquisition of data and recruitment of participants, follow-up, and the definitions and types of chronic pain. Three studies used data from the UK biobank which may mean that some participants are duplicated in our synthesis. To explore the impact of these variations, six subgroup analyses were conducted in the exploratory meta-analyses, including sample size, data source, pain definition, study location, study start date, and publication date. However, the subgroup analyses did not reveal a variable that contributed to the heterogeneity observed. There may be other factors that contributed to the variation, such as the recording of the causes of deaths and reporting bias within individual studies. Finally, some studies did not report raw data on the number of deaths and the age at the time of death, limiting the ability to calculate the impact of chronic pain on life expectancy. Therefore, standardised outcome measures for investigating mortality in chronic pain are needed for observational studies to improve evidence synthesis.

### 4.3 Implications for practice, policy, and future research

To improve the evidence base and ability for evidence synthesis, primary research on mortality in people with chronic pain should report the following variables: mean age at diagnosis and death, causes of death and definitions of pain using ICD-11 codes, pre-existing health conditions, and lifestyle and social factors. This information would help determine the causation and directionality of the risk factors reported in this systematic review. Sharing raw anonymised data, as provided by 84% of included studies, would enhance the ability to synthesise the evidence. Research should be undertaken to determine the most appropriate metric for mortality rates, such as MRR, to standardise the reporting of primary outcomes.

The risk factors for death identified in our review including smoking, low physical activity, and opioid use should be addressed when managing people with chronic pain. Our findings that people with chronic pain have a 30% increased risk of death compared to those without pain is an important outcome to discuss with patients. A study into patient and physician goals of pain management showed that each party has different priorities for treatment [17]. Patients want to focus on pain reduction, while clinicians want to improve function and reduce analgesic prescriptions. Managing differing expectations and ensuring patients are aware of the increased risks of mortality and medications such as opioids is important for improving health outcomes and shared decision-making. However, it is unclear whether people with chronic pain would want to be informed regarding this increased risk of death.

The growing burden of chronic pain may also be contributing to the increase in excess mortality, which has significant policy implications. Our findings illustrate the importance of reporting chronic pain as a contributory factor to death. As the new ICD-11 codes are phased in, doctors and healthcare professionals should be trained in how to accurately document the new codes to improve data validity.

## 5 Conclusions

People with chronic pain have a higher risk of mortality than the general population, which is likely driven by the burden of the condition, opioid use, and lifestyle and social factors such as smoking and low physical activity. Primary outcomes should be standardized, and raw data should be shared to facilitate evidence synthesis. Collaboration between patients, clinicians, and policymakers is needed to optimise interventions such as smoking cessation, physical activity programmes, and alternative therapies to opioids to reduce mortality in people with CNCP.

## Data Availability

All data produced are available online on OSF. See link.

https://osf.io/cp3fx/?view_only=09efe9b9b60e4d81950f248cb08f6867

## 6 Acknowledgements

SW designed the search strategy, performed searches, and screened all articles at all stages. SW extracted and analysed all data, performed the quality assessment, and co-authored this systematic review with GCR. GCR designed the project with SW, checked data extraction, and co-authored the manuscript. AR and LS performed the second screening for both the title/abstract and full-text stages.

## 7 Conflict of Interest

SW received funding from the Wellcome Trust to study for a Doctor of Philosophy (PhD) from 2017-2022, this systematic review was submitted by SW to satisfy in part the coursework requirements for the graduate-entry medicine course at the University of Oxford in the academic year 2022/2023, has not been previously published, and is now eligible for submission external from the university. GCR is the Director of a limited company that is independently contracted to work as an Epidemiologist and teach at the University of Oxford. GCR received scholarships from NIHR SPCR, the Naji Foundation, and the Rotary Foundation between 2017-2021 to study for a Doctor of Philosophy (DPhil/PhD). AOR and LS have no conflicts of interest. No funding has been obtained to conduct this review.

## 11 Supplemental Figures and Tables

**Table S1:** Embase 1974 to present search conducted.

| Embase 1974 to present | Search terms | No. Hits |
| --- | --- | --- |
| 1 | chronic<br>pain*.ab,ti. | 70596 |
| 2 | chronic pain.sh. | 76246 |
| 3 | chronic non<br>cancer<br>pain*.ab,sh,ti. | 1464 |
| 4 | chronic<br>noncancer<br>pain.ab,ti. | 1035 |
| 5 | (chronic adj3<br>pain).ab,ti. | 117429 |
| 6 | primary<br>pain*.ab,ti. | 524 |
| 7 | secondary<br>pain*.ab,ti. | 191 |
| 8 | mortality.ab,ti. | 1429274 |
| 9 | mortality.sh. | 888959 |
| 10 | mortalit*.ab,ti. | 1440267 |
| 11 | death*.ab,ti. | 1450782 |
| 12 | death*.sh. | 318536 |
| 13 | fatalit*.ab,ti. | 45844 |
| 14 | fatality.ab,ti. | 31229 |
| 15 | 1 or 2 or 3 or 4 or<br>5 or 6 or 7 | 137919 |
| 16 | 8 or 9 or 10 or 11<br>or 12 or 13 or 14 | 2844757 |
| 17 | 15 and 16 | 4837 |

**Table S2:** Medline 1946 to present search conducted.

| Medline (Ovid MEDLINE® Epub Ahead of Print, In-Process & Other<br>Non-Indexed Citations, Ovid MEDLINE® Daily and Ovid MEDLINE®)<br>1946 to present | Search terms | No. Hits |
| --- | --- | --- |
| 1 | chronic pain*.ab,ti. | 46685 |
| 2 | chronic pain.sh. | 21730 |
| 3 | chronic non cancer<br>pain*.ab,sh,ti. | 839 |
| 4 | chronic noncancer<br>pain.ab,ti. | 827 |
| 5 | (chronic adj3<br>pain).ab,ti. | 78118 |
| 6 | primary pain*.ab,ti. | 334 |
| 7 | secondary pain*.ab,ti. | 131 |
| 8 | mortality.ab,ti. | 949037 |
| 9 | mortality.sh. | 49267 |
| 10 | mortalit*.ab,ti. | 956214 |
| 11 | death*.ab,ti. | 990629 |
| 12 | death*.sh. | 56358 |
| 13 | fatalit*.ab,ti. | 35952 |
| 14 | fatality.ab,ti. | 24375 |
| 15 | 1 or 2 or 3 or 4 or 5 or<br>6 or 7 | 82713 |
| 16 | 8 or 9 or 10 or 11 or<br>12 or 13 or 14 | 1782831 |
| 17 | 15 and 16 | 2292 |

**Table S3:** Crude death data available from included studies.

| **Study ID (name of first author and year of publication)** | **Population with chronic pain (n)** | **Number of deaths in the population with chronic pain (n)** | **Population no chronic pain (n)** | **Number of deaths in the population with no chronic pain (n)** |
| --- | --- | --- | --- | --- |
| *Andersson et al. 2009* | 888 | 189 | 721 | 67 |
| *Andersen et al. 2016* | 13084 | 3182 | 17309 | 5693 |
| *Chen et al. 2021* | 187269 | 14040 | 197098 | 11877 |
| *Dreyer et al. 2010* | 1189 | 29 | Not reported | Not reported |
| *Ekholm et al. 2014* | 2557 | 426 | 10570 | 769 |
| *Elliott et al. 2014* | 3739 | 952 | Not reported | Not reported |
| *Inoue et al. 2022* | 2168 | 216 | 11710 | 999 |
| *Kim et al. 2019* | 1231 | 256 | 6153 | 1343 |
| *Macfarlane et al. 2001* | 1005 | 129 | 2388 | 196 |
| *Macfarlane et al. 2017* | 7130 | 310 | 275225 | 5269 |
| *Macfarlane et al. 2020* | 201827 | 7900 | 262589 | 8417 |
| *Maruta et al. 1998* | 249 | 22 | Not reported | Not reported |
| *McBeth et al. 2009* | 761 | 217 | 1993 | 389 |
| *Nitter et al. 2013* | 1168 | 57 | 870 | 14 |
| *Vaegter et al. 2018* | 6142 | 276 | Not reported | Not reported |
| *Wolfe et al. 2010* | 8186 | 539 | Not reported | Not reported |

**Table S4:** Causes of death reported by studies.

| **Causes of death** | **Chronic noncancer pain**<br>**(CNCP)** | **non pain** |
| --- | --- | --- |
| *Malignancy* | 8511 | 6533 |
| *Cardiovascular disease* | 4679 | 2274 |
| *Nervous system disease* | 764 | 649 |
| *Mental health disorders* | 317 | 239 |
| *Endocrine and metabolic disease* | 186 | 109 |
| *Respiratory disease* | 166 | 720 |
| *Suicide* | 137 | 2 |
| *Genitourinary disease* | 97 | 62 |
| *Falls* | 90 | 0 |
| *Musculoskeletal and connective*<br>*tissue disease* | 66 | 34 |
| *Accidents* | 47 | 0 |
| *Septicaemia* | 24 | 0 |
| *Kidney Disease* | 12 | 0 |
| *Liver and biliary disease* | 10 | 0 |
| *Gastrointestinal disease* | 1 | 382 |
| *Overdose* | 1 | 0 |
| **Total** | 15108 | 11004 |

**Table S5:** Quality assessment of included studies.

| Study ID (First Author, year) | 1 | 2 | 3 | 4 | 5 | 6 | 7 | 8 | 9 | 10 | 11 | 12 | 13 | 14 | Rating |
| --- | --- | --- | --- | --- | --- | --- | --- | --- | --- | --- | --- | --- | --- | --- | --- |
| Andersson et al. 2009 | ✓ | ✓ | ✓ | ✓ | ✓ | ✓ | ✓ | ✓ | ✓ | X | ✓ | X | ✓ | ✓ | Good |
| Andorsen et al. 2016 | ✓ | ✓ | ✓ | ✓ | ✓ | ✓ | ✓ | ✓ | ✓ | X | ✓ | X | X | ✓ | Fair |
| Chen et al. 2021 | ✓ | ✓ | ✓ | ✓ | ✓ | ✓ | ✓ | ✓ | ✓ | X | ✓ | N/A | N/A | ✓ | Good |
| Dreyer et al. 2010 | ✓ | ✓ | ✓ | ✓ | ✓ | ✓ | ✓ | X | ✓ | X | ✓ | ✓ | ✓ | X | Fair |
| Ekholm et al. 2014 | ✓ | ✓ | N/A | ✓ | ✓ | ✓ | ✓ | X | ✓ | X | ✓ | X | N/A | ✓ | Good |
| Elliott et al. 2013 | ✓ | ✓ | ✓ | ✓ | ✓ | ✓ | ✓ | X | ✓ | X | ✓ | X | ✓ | ✓ | Good |
| Inoue et al. 2022 | ✓ | ✓ | ✓ | ✓ | ✓ | ✓ | ✓ | X | ✓ | X | ✓ | X | ✓ | ✓ | Good |
| Kim et al. 2019 | ✓ | ✓ | ✓ | ✓ | ✓ | ✓ | ✓ | ✓ | ✓ | X | ✓ | X | N/A | ✓ | Good |
| Macfarlane et al. 2001 | ✓ | ✓ | ✓ | ✓ | ✓ | ✓ | ✓ | ✓ | ✓ | X | ✓ | X | N/A | ✓ | Good |
| Macfarlane et al. 2017 | ✓ | ✓ | X | ✓ | ✓ | ✓ | ✓ | X | ✓ | X | ✓ | X | N/A | ✓ | Good |
| Macfarlane et al. 2020 | ✓ | ✓ | ✓ | ✓ | ✓ | ✓ | ✓ | ✓ | ✓ | X | ✓ | X | N/A | ✓ | Good |
| Maruta et al. 1998 | ✓ | ✓ | ✓ | ✓ | X | ✓ | ✓ | X | ✓ | X | ✓ | X | ✓ | X | Fair |
| McBeth et al. 2009 | ✓ | ✓ | ✓ | ✓ | ✓ | ✓ | ✓ | X | ✓ | X | ✓ | X | ✓ | ✓ | Good |
| Nitter et al. 2012 | ✓ | ✓ | ✓ | ✓ | ✓ | ✓ | ✓ | ✓ | ✓ | X | ✓ | X | ✓ | ✓ | Good |
| Sjögren et al. 2010 | ✓ | ✓ | X | ✓ | ✓ | ✓ | ✓ | X | ✓ | X | ✓ | N/A | X | ✓ | Fair |
| Smith et al. 2003 | ✓ | ✓ | ✓ | ✓ | ✓ | ✓ | ✓ | X | ✓ | X | ✓ | X | N/A | ✓ | Good |
| Vaegter et al. 2018 | ✓ | ✓ | ✓ | ✓ | ✓ | ✓ | X | X | ✓ | X | ✓ | ✓ | ✓ | X | Fair |
| Wang et al. 2020 | ✓ | ✓ | N/A | ✓ | ✓ | ✓ | ✓ | X | ✓ | N/A | ✓ | N/A | N/A | ✓ | Good |
| Wolfe et al. 2010 | ✓ | ✓ | ✓ | ✓ | ✓ | ✓ | ✓ | X | ✓ | X | ✓ | X | ✓ | ✓ | Good |

**Figure S6:**
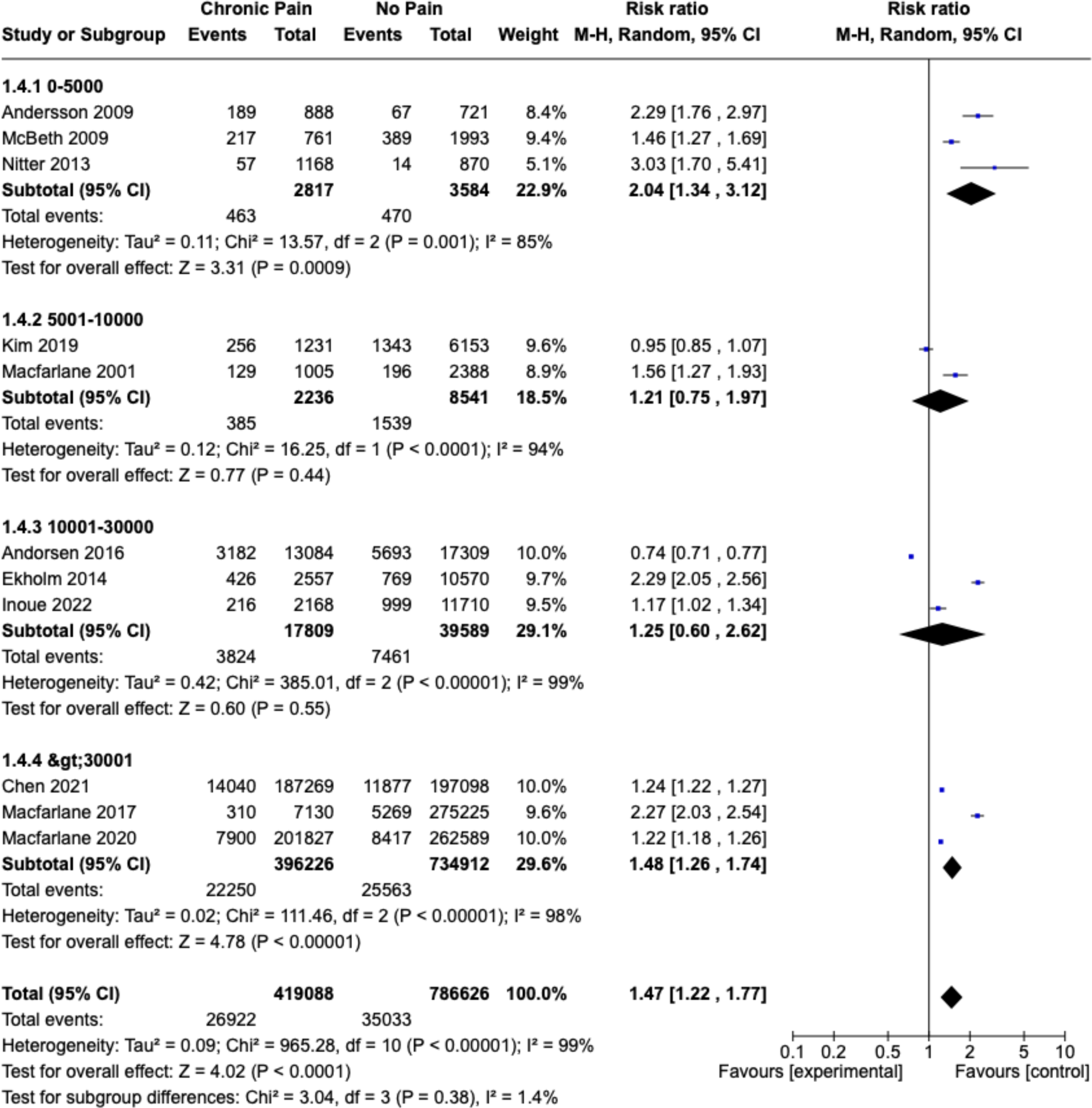
Sensitivity analysis of sample size.

**Figure S7:**
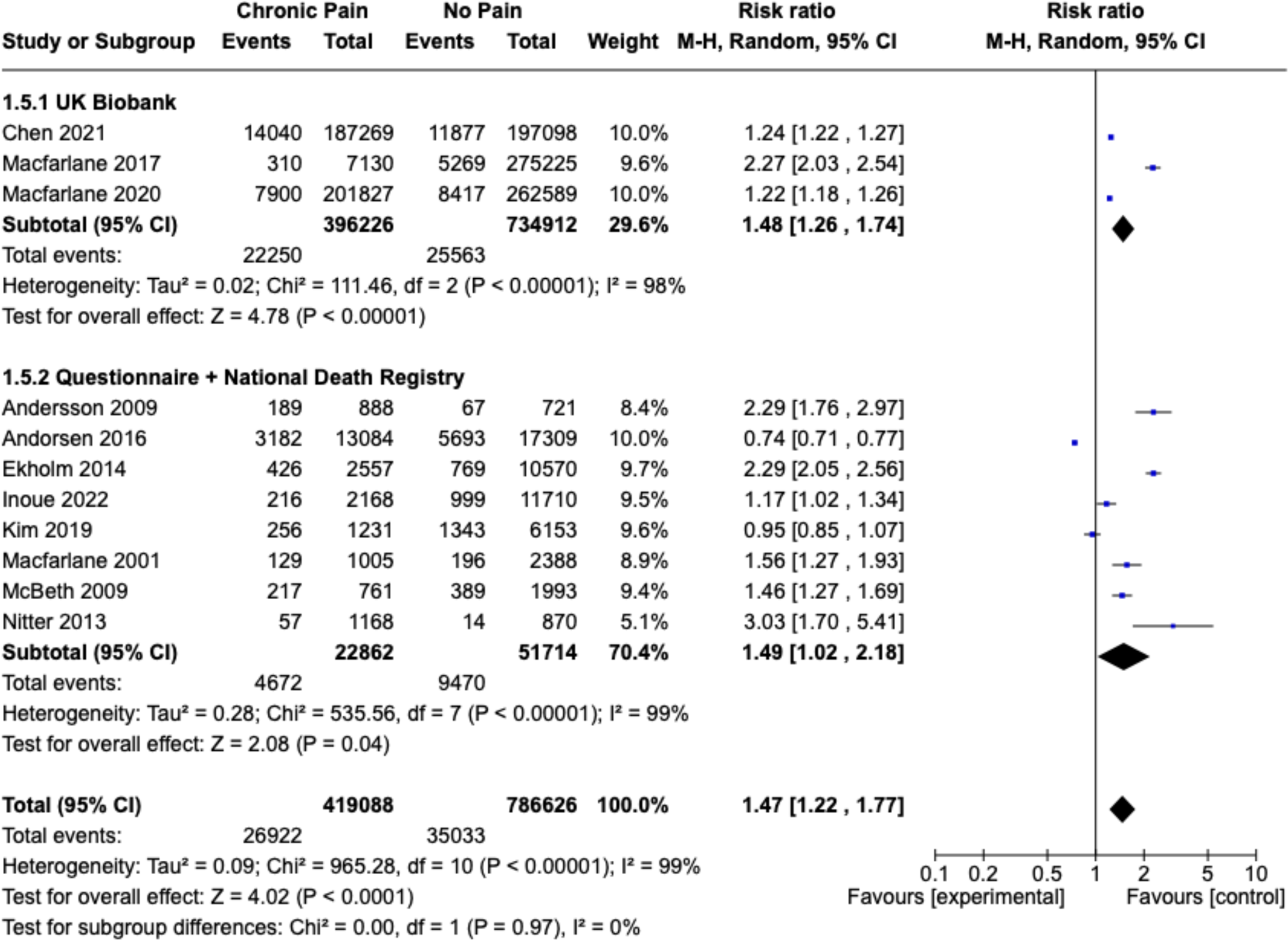
Sensitivity analysis of data sources.

**Figure S8:**
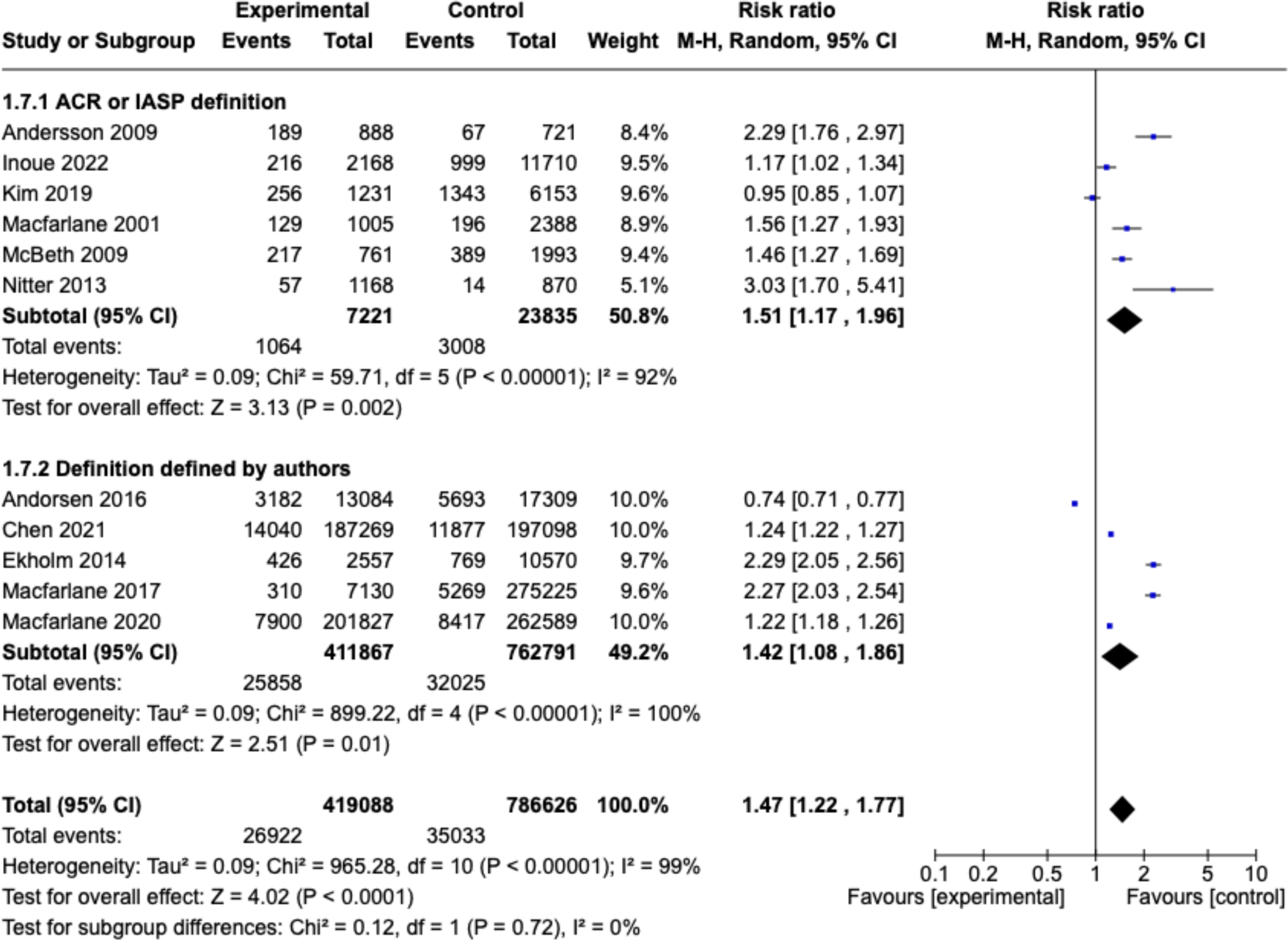
Sensitivity analysis by pain definition.

**Figure S9:**
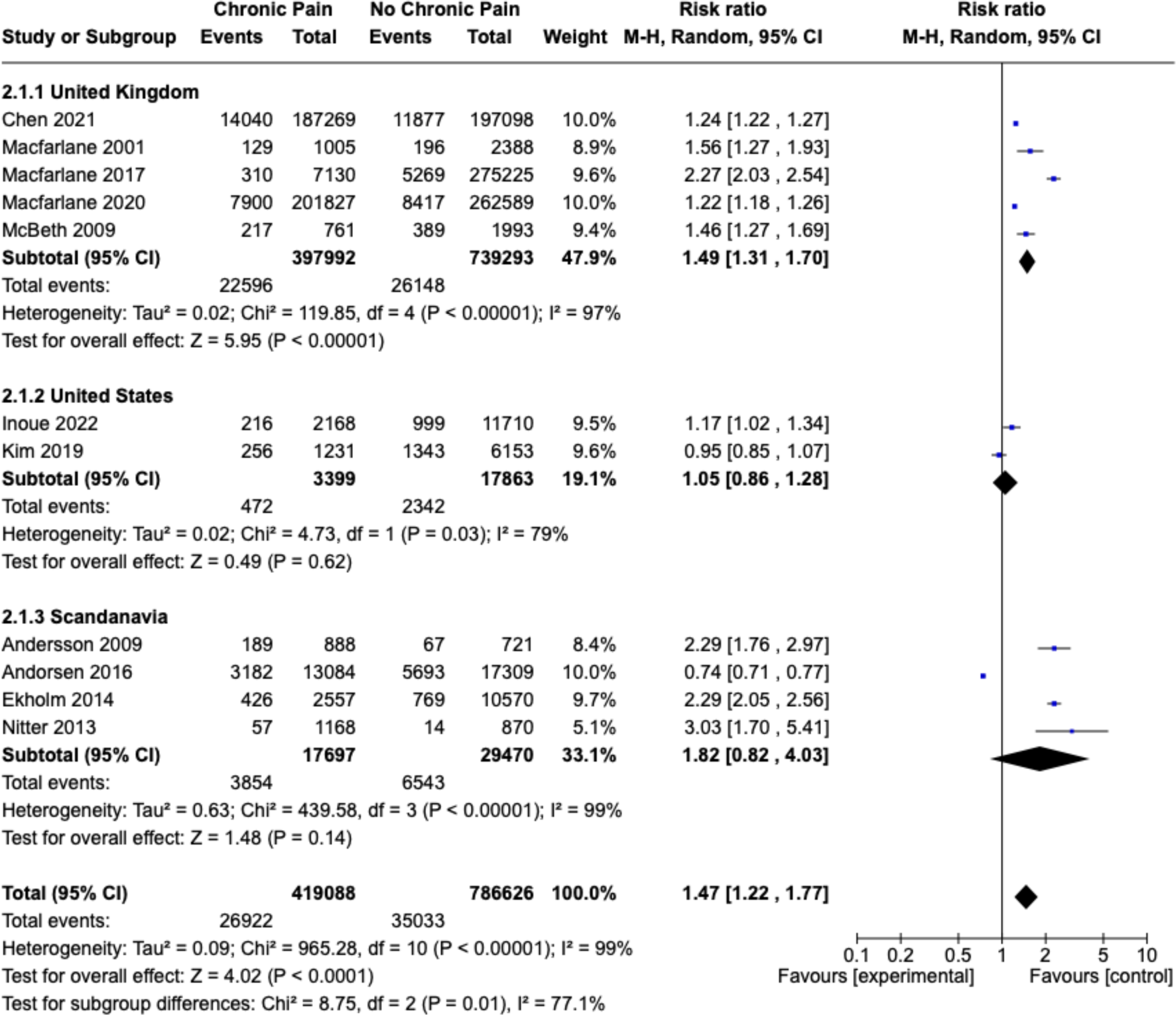
Sensitivity analysis by study location.

**Figure S10:**
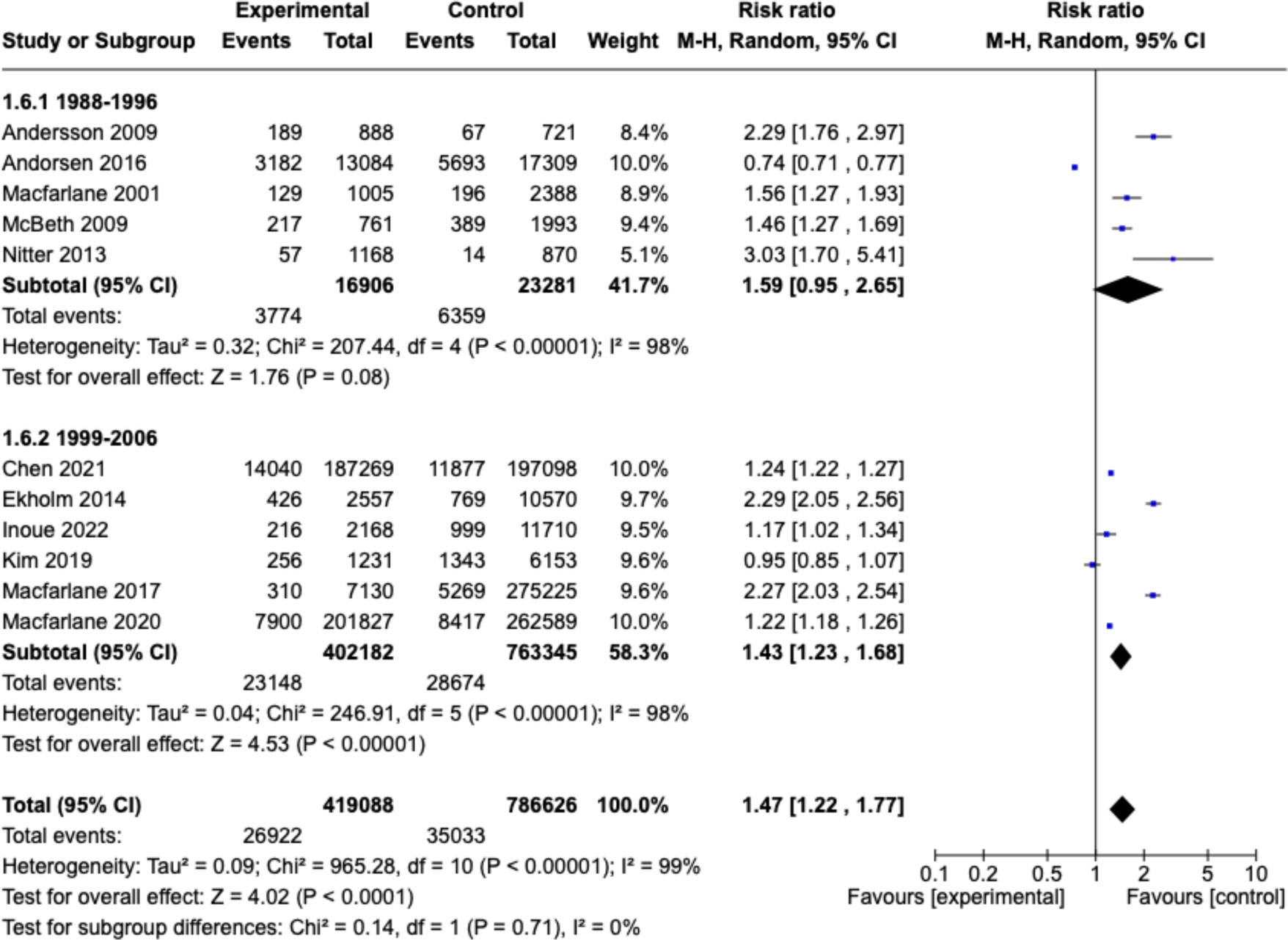
Sensitivity analysis by study start date.

**Figure S11:**
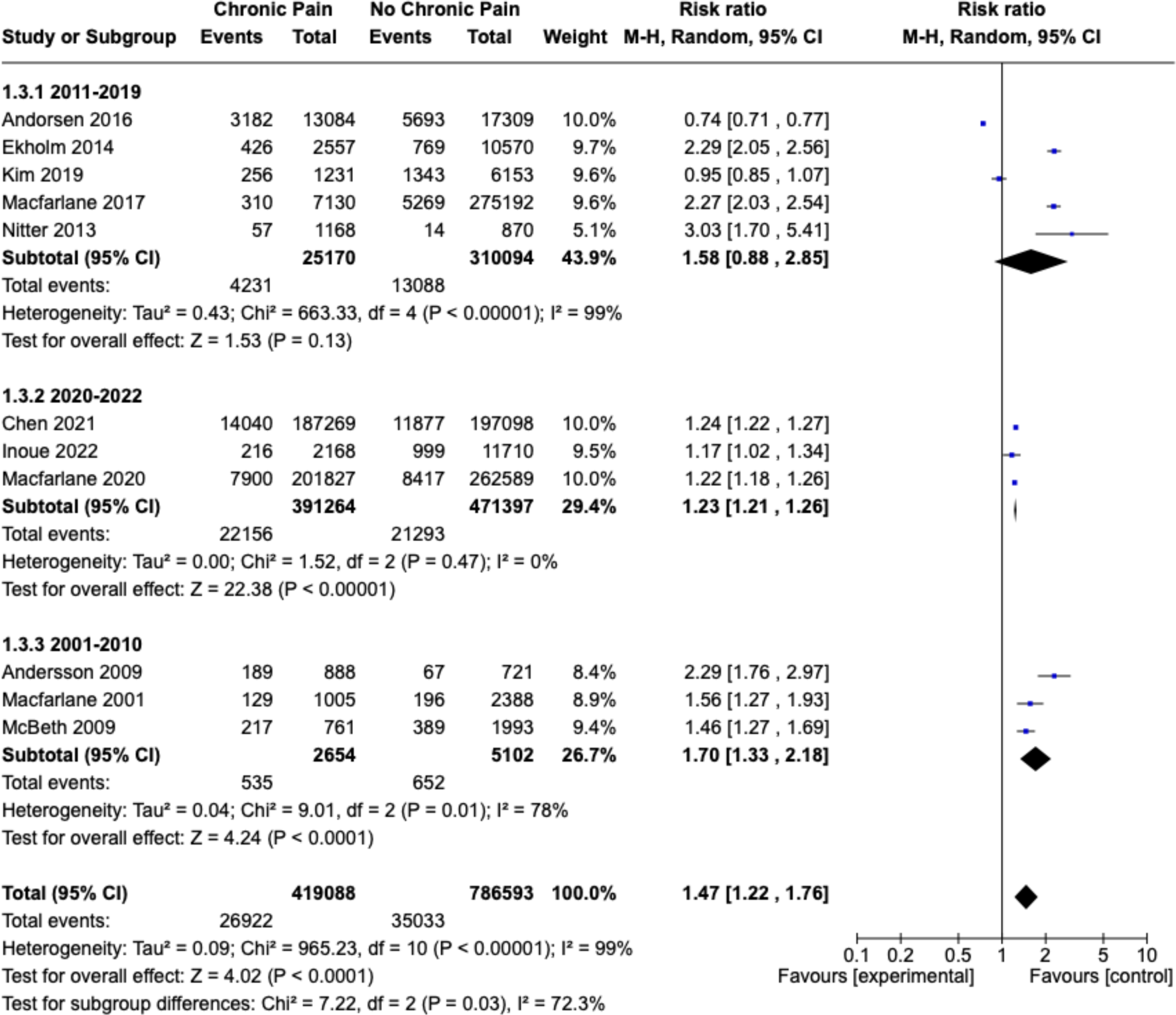
Sensitivity analysis by study publication date.

## Notes

### Competing Interest Statement

The authors have declared no competing interest.

### Clinical Protocols

https://osf.io/cp3fx/?view_only=09efe9b9b60e4d81950f248cb08f6867

### Funding Statement

This study did not receive funding.

